# Population-based seroprevalence of SARS-CoV-2 is more than halfway through the herd immunity threshold in the State of Maranhão, Brazil

**DOI:** 10.1101/2020.08.28.20180463

**Authors:** Antônio Augusto Moura da Silva, Lídio Gonçalves Lima-Neto, Conceição de Maria Pedrozo e Silva de Azevedo, Léa Márcia Melo da Costa, Maylla Luana Barbosa Martins Bragança, Allan Kardec Duailibe Barros Filho, Bernardo Bastos Wittlin, Bruno Feres Souza, Bruno Luciano Carneiro Alves de Oliveira, Carolina Abreu de Carvalho, Erika Barbara Abreu Fonseca Thomaz, Eudes Alves Simões-Neto, Jamesson Ferreira Leite Júnior, Lécia Maria Sousa Santos Cosme, Marcos Adriano Garcia Campos, Rejane Christine de Sousa Queiroz, Sérgio Souza Costa, Vitória Abreu de Carvalho, Vanda Maria Ferreira Simões, Maria Teresa Seabra Soares de Brito e Alves, Alcione Miranda dos Santos

## Abstract

**Background:** Few population-based studies on the prevalence of severe acute respiratory syndrome coronavirus 2 (SARS-CoV-2) have been performed to date, and most of them have used lateral flow immunoassays with finger-prick, which may yield false-negative results and thus underestimate the true infection rate.

**Methods:** A population-based household survey was performed in the State of Maranhão, Brazil, from 27 July 2020 to 8 August 2020 to estimate the seroprevalence of SARS-CoV-2 using a serum testing electrochemiluminescence immunoassay. A three-stage cluster sampling stratified by four state regions was used. The estimates took clustering, stratification, and non-response into account. Qualitative detection of IgM and IgG antibodies was performed in a fully-automated Elecsys® Anti-SARS-CoV-2 electrochemiluminescence immunoassay on the Cobas® e601 analyser (Roche Diagnostics).

**Findings:** A total of 3156 individuals were interviewed. Seroprevalence of total antibodies against SARS-CoV-2 was 40·4% (95%CI 35·6-45·3). Population adherence to non-pharmaceutical interventions was higher at the beginning of the pandemic than in the last month. SARS-CoV-2 infection rates were significantly lower among mask wearers and among those who maintained social and physical distancing in the last month compared to their counterparts. Among the infected, 62·2% had more than three symptoms, 11·1% had one or two symptoms, and 26·0% were asymptomatic. The infection fatality rate was 0·17%, higher for males and advanced age groups. The ratio of estimated infections to reported cases was 22·2.

**Interpretation:** To the best of our knowledge, the seroprevalence of SARS-CoV-2 estimated in this population-based survey was the highest and the closest to the herd immunity threshold reported to date. Our results suggest that the herd immunity threshold is not as low as 20%, but at least higher than or equal to around 40%. The infection fatality rate was one of the lowest reported so far, and the proportion of asymptomatic cases was low.

## Introduction

Of all the countries, Brazil is one of the most severely affected by coronavirus disease 2019 (COVID-19) pandemic. Its first case was reported on 26 February 2020, and by 20 August 2020 3,501,975 cases were reported, with 112,304 deaths, the second-highest in the world.^1^ The national response has been controversial, testing capacity is low, and disagreements among the different levels of government over social distancing measures conveyed contradictory messages to the population. As a middle-income country, Brazil has high poverty rates and an extensive part of its population is engaged in informal activities that face difficulties to make ends meet and to follow stay-at-home measures.^2^ As a consequence of all these facts, social distancing has not reached levels sufficient to curb and contain the COVID-19 pandemic.^3^

The State of Maranhão is located in the Northeast region of Brazil and has an estimated population of 7,114,598 inhabitants in 2020^4^ with an area of 329,642 km², a little larger than that of Italy. It is one of the states in Brazil where the pandemic gathered speed early. Its first case was reported on 20 March 2020, and by 20 August 2020, the number of deaths reported was 3315. Deaths peaked in May and decreased thereafter. From 3 May 2020 to 17 May 2020 São Luís Island, where the state capital city is located, was put into lockdown. Reduction of social mobility reached at most 55% at the end of March and during lockdown at the capital but remained low (40%-45%) during the worst phase of the pandemic. Despite low home quarantine adherence, the number of deaths decreased, and intensive care units occupancy diminished.^5^

Herd immunity to severe acute respiratory syndrome coronavirus 2 (SARS-CoV-2) is an ongoing debate. Although many consider it to be around 60%-70%, using the classical formula 1-1/R^0^, where R^0^ is the basic reproductive number,^6^ some reports have proposed that herd immunity could be as low as 10%-20%^7^ or around 43%,^8^ due to the heterogeneity in susceptibility or exposure to infection across population groups.^7,8^ However, reported population-based seroprevalences of SARS-CoV-2 were lower than the herd immunity thresholds, ranging from extremely low infections rates, close to 1%-3%,^9,10^ to values as high as 14·3% in Barcelona,^11^ Spain, and 22·7% in New York City.^12^ In Brazil, the highest reported population-based seroprevalence was 17·9%, for the São Paulo municipality.^13^

The infection fatality rate (IFR) and the percentage of asymptomatic infections of SARS-CoV-2 are known with uncertainty. Early reports at the beginning of the pandemic estimated IFR at values between 0·6% and 1·3%,^14,15^ and considered asymptomatic infections as being highly prevalent.^14,16^ Most recent reviews, however, estimated a lower IFR with large variations across sites^10,17^ and a much lower percentage of asymptomatic infections.^11,18,19^

Population-based surveys are necessary to monitor the infection progression since most cases are undocumented.^20^ However, few population-based studies on the prevalence of SARS-CoV-2 have been performed to date, and most of them have used lateral flow immunoassays with finger-prick, which may yield false-negative results, and thus underestimate the true infection rate.^21,22^ Therefore, population-based surveys using more sensitive diagnostic tests are warranted. In this population-based study, we estimated the overall seroprevalence of SARS-CoV-2 using a serum testing electrochemiluminescence immunoassay. Sociodemographic characteristics of the population, self-reported symptoms, adherence to non-pharmaceutical interventions (NPIs), use of health services, previous molecular and antibody testing among the infected, and the IFR were also assessed.

## Methods

### Study design and participants

A cross-sectional survey to estimate the seroprevalence of antibodies against SARS-CoV-2 was conducted from 27 July 2020 to 8 August 2020 by population-based household sampling. The study was developed in cooperation between the Federal University of Maranhão and the State Health Department of Maranhão, Brazil.

Conglomerate sampling in three stratified stages in four regions was used. The regions were the Island of São Luís including the state capital, small municipalities (<20,000 inhabitants), medium-sized municipalities (20,000 to 100,000 inhabitants) and large municipalities except for the island (>100,000 inhabitants). In each stratum, in the first stage, 30 census tracts were selected by systematic sampling. In the second stage, 34 households were selected in each census tract by systematic sampling. In the third stage, an eligible resident (residing for at least six months in the household) aged one year or more was selected by simple random sampling using a table of random numbers.

### Data collection, instruments, and variables

Trained professionals from the municipal and state health departments were responsible for data collection. Using a map provided by the Brazilian Institute of Geography and Statistics (IBGE in the Portuguese acronym) the starting point (identified with an X on the map) and the geographic boundaries of each census tracts were identified. The first interview was held in the home closest to the starting point of each sector. Then, facing that domicile, the interviewer walked to the left with his/her right shoulder facing the wall/residences. Without including the visited home, the interviewer counted five residences and conducted the next interview in the fifth one. If the selected household was empty at the time or the elected person did not agree to participate in the survey, the next house to the left (neighbour) of the original one was taken as a replacement. If that house was also empty or if the elected person refused to participate the next house to the left was visited. Then the interviewer counted five domiciles and conducted the next interview in the fifth house after the original one. The team always proceeded to the left in relation to the last surveyed domicile and conducted the next interview in the fifth domicile. Non-residential buildings were excluded from the count. After completing the tour on the block, the interviewer facing the last visited domicile continued to the next adjacent block located to the left, always adopting the same strategy.

A questionnaire with closed-ended questions was applied in a face-to-face interview with the individual or his/her legal guardian. The questionnaire was composed of sociodemographic questions, adherence to NPIs, self-reported symptoms, and the use of health services. The sociodemographic questions included sex, age group, skin colour/race, head of the household’s schooling, monthly family income in Brazilian Reals, and the number of the household residents. Head of the household’s schooling was classified according to the International Standard Classification of Education (ISCED) 2011 into early childhood/primary/lower secondary education (levels 0-2), upper secondary/post-secondary non-tertiary education (levels 3-4), and tertiary education and beyond (levels 5-8).^23^ Skin colour/race was categorized according to the IBGE and divided into white, brown, or black.^24^ Yellows and indigenous were excluded because they were too few for a meaningful analysis.

Adherence to NPI at the beginning of the pandemic and in the last month included social distancing (yes, if the person never leaves home or seldom goes out, with a maximum of one outing every fifteen days, and no otherwise), wearing of face masks (yes, if the individual uses a mask on all exits and does not remove or seldom removes the mask from the face, and no otherwise), hand hygiene (yes, if the person sanitizes the hands more than six times per turn with soap or an alcohol gel, and no otherwise), and physical distancing (yes, if the individual never or hardly ever comes within 1·5 m of other people, and no otherwise).

A self-reported symptom rating, adapted from Pollán et al. (2020),^11^ was used and the persons were classified into asymptomatic; oligosymptomatic: the presence of one to two symptoms without anosmia/hyposmia or ageusia/dysgeusia; and symptomatic: anosmia/hyposmia or ageusia/dysgeusia or more than two symptoms including fever, chills, sore throat, cough, dyspnoea, diarrhoea, nausea/vomiting, headache, fatigue, and myalgia.

Questions on the use of health services included if the individual looked for health services, received care when seeking health services, was hospitalized for over 24 hours, received a medical diagnosis of suspected COVID-19, performed RT-PCR for SARS-CoV-2, and performed an antibody test– point-of-care/serology for SARS-CoV-2.

Data were abstracted into the *Epicollect5 Data Collection* mobile application.

### SARS-CoV-2 antibodies detection

For the qualitative determination of antibodies against SARS-CoV-2, 5·0 ml of whole blood was collected, and after centrifugation at 1800 g for 15 min, the serum was obtained. Then, a highly sensitive and specific sandwich electrochemiluminescence immunoassay (Elecsys® Anti-SARS-CoV-2 assay, Roche Diagnostics) was used to detect IgM and IgG antibodies against the SARS-CoV-2 nucleocapsid antigen according to the manufacturer’s instruction using a fully automated Cobas® e601 immunoassay analyser (Roche Diagnostics).^25^

### Sample size calculation

The formula used to determine the sample size in each stratum was given by

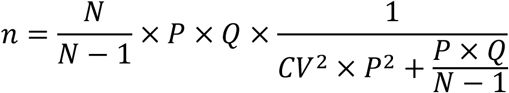

N being the population size in each stratum; P the expected prevalence in the stratum; Q=1-P; and CV the coefficient of variation of the prevalence estimates within the stratum. In each stratum, the expected prevalence of infection was 20%, and the coefficient of variation was 10%. For the final calculation, a design effect of 2 was added. Thus, the minimum number of individuals per stratum was 800, totalling 3200 individuals to compose the sample. Predicting losses, the sample size was increased by 25% resulting in 4000 observations.

### Statistical analysis

The basic sample weight of each selected unit (census sector, household, and individual) was calculated separately for each stratum, considering the inverse of the selection probability according to the sampling plan specified for the study.

The probability of selection of the census sector “j” in each stratum “i” of the sample is given by 30/S_i_, where “S_i_” is the number of census sectors of the stratum “i” in the population and the probability of the domicile of the census sector “j” of the stratum “i” being selected was obtained from the following expression: 34/D_ij_, where “D_ij_” is the number of domiciles in sector “j” of the stratum “i” in the population. The probability of each resident in the selected household was given by 1/(number of residents in the household). The number of sectors and domiciles was obtained from the 2010 Census of the IBGE.

Since losses, refusals, and non-responses occurred, the response rate in each stratum was also calculated. Considering that there were three stages, the final weight was obtained by the product of the basic weight in each stage and the response rate.

All analyses were performed using R version 4.0.2. Weighting factors, clustering, and stratification were incorporated into the analyses via the R survey package. Prevalence and 95% confidence interval (95%CI) of SARS-CoV-2 infection was obtained according to the sociodemographic characteristics, adherence to NPI, self-reported symptoms, and the use of health services. The chi-square test, considering the study design, was used to compare the prevalence between groups. The McNemar test was used to compare adherence to NPI over time.

The IFR was estimated by dividing the estimated number of deaths by the estimated proportion of infections obtained by the serological survey multiplied by the stratified age and sex population estimates.^4^ The number of deaths reported on 8 August 2020 was abstracted from official sources.^5^ The IFR was estimated by the sex, the sex and age group, and the overall IFR was evaluated for the State of Maranhão and the first stratum (São Luís Island). The number of deaths occurring daily was estimated using Nowcasting by Bayesian Smoothing (NobBS), to consider reporting delays and underreporting of deaths attributable to COVID-19 (assuming a prior of 15% for the State of Maranhão and 10% for the São Luís Island). This procedure incorporates uncertainty both in the delay distribution and in the evolution of the pandemic curve over time, resulting in smooth, time-correlated estimates of the number of deaths.^26^ Simulations were carried out using the NobBS R package, with a negative binomial model with an adaptation phase of 10000 iterations and a burn-in of 10000 iterations for estimating deaths in the State of Maranhão, and the same parameters with 5000 iterations for the São Luís Island. The case reporting rate was obtained by dividing the number of reported cases by the number of estimated infections, multiplied by 100.

### Ethical approval

Ethical approval was obtained from the Research Ethics Committee of the Carlos Macieira Hospital of the Maranhão State Health Secretariat under CAAE number 34708620.2.0000.8907. Written informed consent was provided by the participants or the parents/legal guardians.

### Role of the funding source

The funder facilitated data acquisition and helped in the analysis, interpretation, and writing.

The corresponding author confirms that he had full access to all the data in the study and had the final responsibility for the decision to submit for publication.

## Results

A total of 3289 individuals (80·6%) agreed to participate in the study. After the exclusion of samples with insufficient material or haemolysed samples, and cases in which it was not possible to link the result of the examination with the person, 3156 participants had their blood samples analysed (77·4%). Comparing the sampling with the population distribution (age and sex estimates for 2020), males and people of working age were underrepresented in the sample.

Seroprevalence of total antibodies against SARS-CoV-2 was 40·4% (95%CI 35·6-45·3) in the State of Maranhão, Brazil. Seroprevalence varied by region, from 20·0% in small municipalities with <20,000 inhabitants, reaching 47·6% in medium-sized municipalities from 20,000 to 100,000 inhabitants (P=0·006). Seroprevalence in the São Luís Island, including the capital of the state, was 38·9%. There were no significant differences in the prevalence according to the sex or age group (Table 1).

**Table 1.** Prevalence of antibodies against SARS-CoV-2 by region, sex, and age group, State of Maranhão, Brazil, 2020.

| Variables | n | % weighted | Population distribution (%) | f infected | % infected weighted (95% CI) | p-value |
| --- | --- | --- | --- | --- | --- | --- |
| Total | 3156 | 100.0 | 100.0 | 1167 | 40.4 (35.6-45.3) |  |
| Region |  |  |  |  |  | 0.006 |
| São Luís Island including the capital | 737 | 25.5 | 20.2 | 349 | 38.9 (24.5-53.2) |  |
| Municipalities with <20,000 inhabitants | 754 | 20.0 | 21.4 | 215 | 31.0 (24.3-37.8) |  |
| Municipalities with 20,000 to 100,000 inhabitants | 839 | 41.6 | 45.1 | 346 | 47.6 (42.0-53.1) |  |
| Municipalities with >100000 inhabitants | 826 | 12.9 | 13.2 | 257 | 35.2 (26.1-44.3) |  |
| Sex |  |  |  |  |  | 0.134 |
| Male | 1200 | 37.1 | 49.1 | 426 | 37.2 (31.8-42.6) |  |
| Female | 1956 | 62.9 | 50.9 | 741 | 42.4 (36.1-48.6) |  |
| Age group (years) * |  |  |  |  |  | 0.230 |
| 1-9 | 124 | 5.2 | 16.5 | 49 | 42.6 (33.8-51.3) |  |
| 10-19 | 330 | 14.7 | 18.6 | 125 | 43.0 (33.5-52.4) |  |
| 20-29 | 427 | 12.5 | 18.0 | 184 | 49.2 (41.1-57.3) |  |
| 30-39 | 475 | 15.0 | 16.0 | 170 | 44.4 (37.4-51.4) |  |
| 40-49 | 502 | 16.3 | 12.0 | 191 | 32.2 (23.4-41.0) |  |
| 50-59 | 501 | 14.6 | 8.5 | 168 | 39.1 (32.1-46.1) |  |
| 60-69 | 409 | 11.7 | 5.7 | 144 | 40.3 (29.1-51.4) |  |
| ≥70 | 386 | 9.8 | 4.8 | 136 | 34.3 (25.7-42.9) |  |

\* Numbers did not add up to total because of missing values
95% CI: 95% confidence interval; SARS-CoV-2: severe acute respiratory syndrome coronavirus 2

Whites had a lower point prevalence (20·0%) compared with both the browns (41·3%) and blacks (49·2%) but of borderline significance (between 0·05 and 0·10). Persons with tertiary education had a lower prevalence of infection (27·5%) than their counterparts (P=0·011). Although point prevalence was lower among those with a monthly family income above 3000 Brazilian Reals, the difference did not reach a significant level. Infection rates were higher among households with three dwellers (44·9%) (P=0·028) (Table 2).

**Table 2.** Prevalence of antibodies against SARS-CoV-2 by race, schooling, family income, and number of residents, State of Maranhão, Brazil, 2020.

| Variables | n | % weighted | f infected | % infected weighted<br>(95% CI) | p-value |
| --- | --- | --- | --- | --- | --- |
| Skin colour/race* |  |  |  |  | 0.080 |
| White | 590 | 20.0 | 200 | 32.2 (20.1-44.4) |  |
| Brown | 2100 | 67.4 | 767 | 41.3 (37.1-45.4) |  |
| Black | 396 | 12.6 | 177 | 49.1 (42.3-55.9) |  |
| Head of the household's schooling<br>(years)** |  |  |  |  | 0.011 |
| Primary/Lower secondary | 1369 | 37.7 | 487 | 40.9 (35.4-46.4) |  |
| Upper secondary | 1251 | 41.9 | 512 | 46.2 (41.3-51.2) |  |
| Tertiary | 517 | 20.4 | 161 | 27.5 (16.9-38.1) |  |
| Monthly family income (Brazilian Real)<br>*** |  |  |  |  | 0.101 |
| <1000 | 607 | 17.7 | 222 | 40.8 (34.6-46.9) |  |
| 1000 a <2000 | 1405 | 42.5 | 540 | 45.9 (41.0-50.9) |  |
| 2000 a <3000 | 617 | 20.8 | 243 | 42.9 (35.7-50.2) |  |
| >3000 | 493 | 19.0 | 155 | 27.9 (17.0-38.9) |  |
| Number of residents |  |  |  |  | 0.028 |
| 1 | 386 | 3.9 | 122 | 35.4 (27.2-43.6) |  |
| 2 | 840 | 15.7 | 302 | 37.7 (32.0-43.5) |  |
| 3 | 739 | 21.2 | 297 | 44.9 (40.0-49.8) |  |
| 4 | 628 | 28.0 | 234 | 38.8 (29.0-48.5) |  |
| ≥5 | 563 | 31.2 | 212 | 40.9 (33.5-48.3) |  |

\* yellow and indigenous were excluded because they were too few for a meaningful analysis \*\* Numbers did not add up to total because of missing values
\*\*\* 1 Brazilian Real (R\$) is equivalent to approximately US\$ 5.60 US dollars
95% CI: 95% confidence interval; SARS-CoV-2: severe acute respiratory syndrome coronavirus 2

Population adherence to NPI to contain the COVID-19 pandemic were mostly higher at the beginning of the pandemic than in the last month. Social distancing dropped from 52·7% to 37·4% (P<0·001). The percentage of wearing a face mask dropped from 61·4% to 55·5% (P<0·001). Differences in the infection rates between those who maintained social distancing and those who did not were evident both at the beginning of the pandemic (36·4% vs 45·0%, P=0·020) and in the last month (34·0% vs 44·3%, P=0·015). SARS-CoV-2 infection rates were significantly lower in the last month among mask wearers and those who maintained a distance of at least 1·5 m from other people compared to their counterparts (P=0·036 for mask-wearing and P=0·030 for physical distancing) (Table 3).

**Table 3.**
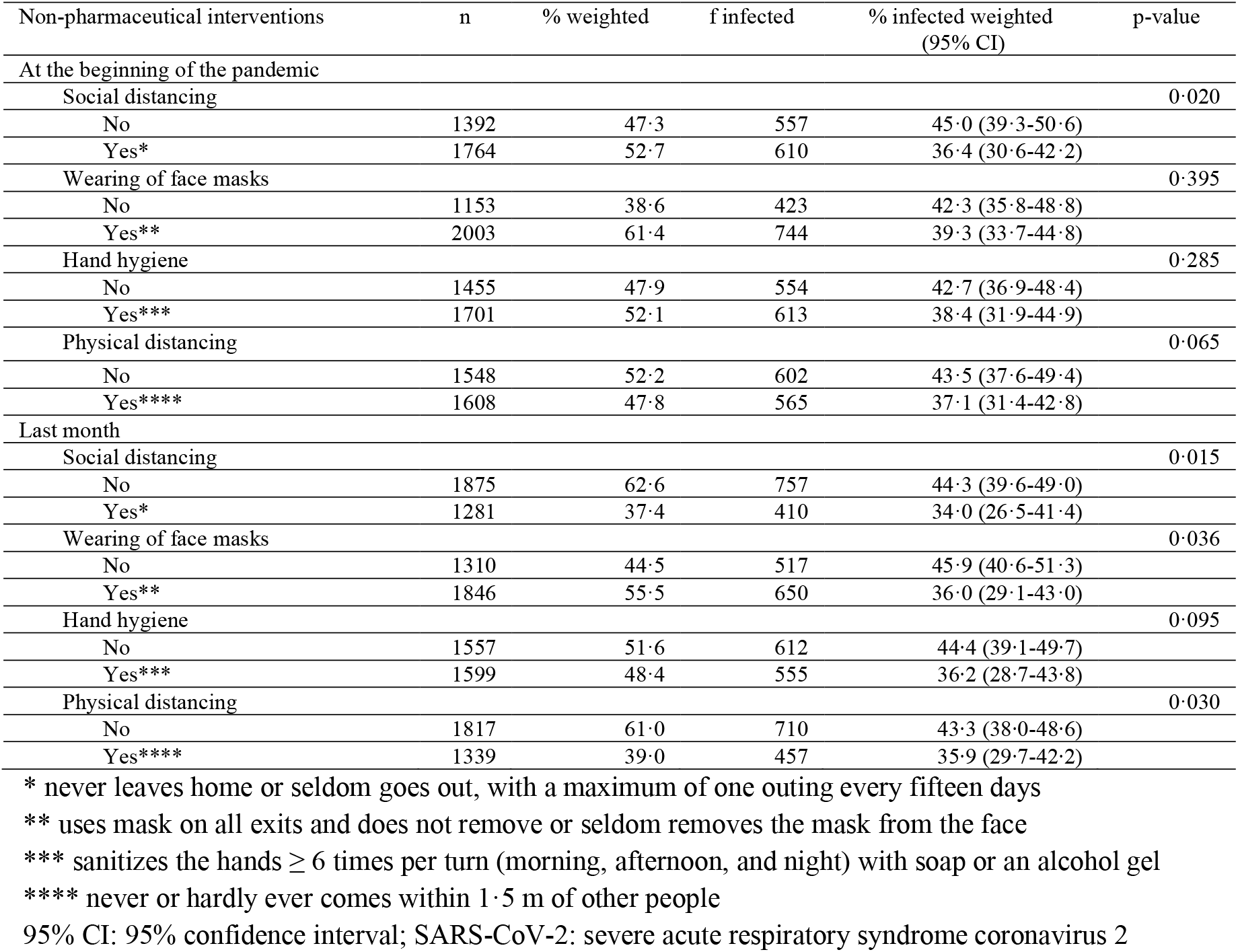
Prevalence of antibodies against SARS-CoV-2 according to adherence to non-pharmaceutical interventions at the beginning of the pandemic and in the last month, State of Maranhão, Brazil, 2020.

| Non-pharmaceutical interventions | n | % weighted | f infected | % infected weighted (95% CI) | p-value |
| --- | --- | --- | --- | --- | --- |
| **At the beginning of the pandemic** |  |  |  |  |  |
| Social distancing |  |  |  |  | 0.020 |
| No | 1392 | 47.3 | 557 | 45.0 (39.3-50.6) |  |
| Yes* | 1764 | 52.7 | 610 | 36.4 (30.6-42.2) |  |
| Wearing of face masks |  |  |  |  | 0.395 |
| No | 1153 | 38.6 | 423 | 42.3 (35.8-48.8) |  |
| Yes** | 2003 | 61.4 | 744 | 39.3 (33.7-44.8) |  |
| Hand hygiene |  |  |  |  | 0.285 |
| No | 1455 | 47.9 | 554 | 42.7 (36.9-48.4) |  |
| Yes*** | 1701 | 52.1 | 613 | 38.4 (31.9-44.9) |  |
| Physical distancing |  |  |  |  | 0.065 |
| No | 1548 | 52.2 | 602 | 43.5 (37.6-49.4) |  |
| Yes**** | 1608 | 47.8 | 565 | 37.1 (31.4-42.8) |  |
| **Last month** |  |  |  |  |  |
| Social distancing |  |  |  |  | 0.015 |
| No | 1875 | 62.6 | 757 | 44.3 (39.6-49.0) |  |
| Yes* | 1281 | 37.4 | 410 | 34.0 (26.5-41.4) |  |
| Wearing of face masks |  |  |  |  | 0.036 |
| No | 1310 | 44.5 | 517 | 45.9 (40.6-51.3) |  |
| Yes** | 1846 | 55.5 | 650 | 36.0 (29.1-43.0) |  |
| Hand hygiene |  |  |  |  | 0.095 |
| No | 1557 | 51.6 | 612 | 44.4 (39.1-49.7) |  |
| Yes*** | 1599 | 48.4 | 555 | 36.2 (28.7-43.8) |  |
| Physical distancing |  |  |  |  | 0.030 |
| No | 1817 | 61.0 | 710 | 43.3 (38.0-48.6) |  |
| Yes**** | 1339 | 39.0 | 457 | 35.9 (29.7-42.2) |  |

\* never leaves home or seldom goes out, with a maximum of one outing every fifteen days
\*\* uses mask on all exits and does not remove or seldom removes the mask from the face
\*\*\* sanitizes the hands $\geq 6$ times per turn (morning, afternoon, and night) with soap or an alcohol gel
\*\*\*\* never or hardly ever comes within 1.5 m of other people
95% CI: 95% confidence interval; SARS-CoV-2: severe acute respiratory syndrome coronavirus 2

Differences in the self-reporting symptoms were highly significant comparing those with and without antibodies to SARS-CoV-2. Among the infected, 62·2% had more than three symptoms, whereas 26·0% were asymptomatic and, 11·8% reported only one or two symptoms (oligosymptomatic). The predominant symptoms among those who tested positive for SARS-CoV-2 were anosmia/hyposmia (49·5%), ageusia/dysgeusia (47·7%), fever (45·6%), headache (45·4%), myalgia (43·6%), and fatigue (41·1%) (Table 4).

**Table 4.** Reported symptoms of SARS-CoV-2 infection, State of Maranhão, Brazil, 2020.

| Variables | Non-infected (n=1989) |  | Infected (n=1167) |  | p-value |
| --- | --- | --- | --- | --- | --- |
|  | n | % weighted<br>(95% CI) | n | % weighted<br>(95% CI) |  |
| Self-reported symptom rating* |  |  |  |  | <0.001 |
| Asymptomatic | 1104 | 52.1 (47.3-56.9) | 320 | 26.0 (21.0-31.0) |  |
| Oligosymptomatic (1 to 2 symptoms) | 427 | 22.8 (18.8-26.7) | 134 | 11.8 (8.9-14.6) |  |
| Symptomatic ($\geq 3$ symptoms) | 458 | 25.2 (21.8-28.5) | 713 | 62.2 (56.2-68.3) | |
| Self-reported symptoms |  |  |  |  |  |
| Fever | 296 | 16.7 (13.1-20.4) | 494 | 45.6 (39.9-51.3) | <0.001 |
| Shivers | 253 | 13.7 (9.9-17.6) | 379 | 34.3 (29.5-39.2) | <0.001 |
| Sore throat | 345 | 18.3 (14.7-22.0) | 378 | 34.5 (30.1-39.0) | <0.001 |
| Cough | 356 | 17.6 (13.7-21.5) | 369 | 33.1 (29.7-36.5) | <0.001 |
| Dyspnoea | 184 | 10.9 (8.4-13.4) | 209 | 18.6 (15.0-22.2) | 0.001 |
| Runny nose | 370 | 19.2 (16.0-22.3) | 364 | 32.2 (28.0-36.5) | <0.001 |
| Palpitations | 204 | 9.5 (7.1-11.9) | 178 | 15.6 (12.1-19.2) | <0.001 |
| Anosmia/Hyposmia | 117 | 7.3 (5.0-9.6) | 547 | 49.5 (42.5-56.5) | <0.001 |
| Ageusia/Dysgeusia | 133 | 8.2 (5.8-10.5) | 535 | 47.7 (40.4-54.9) | <0.001 |
| Diarrhoea | 186 | 8.9 (6.5-11.2) | 210 | 18.1 (15.1-21.2) | <0.001 |
| Nausea/vomiting | 146 | 7.2 (5.2-9.2) | 177 | 15.1 (12.0-18.2) | <0.001 |
| Headache | 509 | 27.5 (22.7-32.2) | 491 | 45.4 (38.7-52.0) | <0.001 |
| Abdominal pain | 201 | 10.8 (7.7-13.9) | 196 | 19.8 (14.5-25.1) | 0.009 |
| Myalgia | 368 | 17.9 (14.0-21.8) | 485 | 43.6 (36.8-50.3) | <0.001 |
| Fatigue | 333 | 17.1 (13.0-21.3) | 449 | 41.1 (34.5-47.7) | <0.001 |
| Loss of appetite | 217 | 11.1 (8.2-14.0) | 396 | 35.2 (29.2-41.2) | <0.001 |

\* Asymptomatic: no symptoms; oligosymptomatic: presence of 1 to 2 symptoms without anosmia/hyposmia or ageusia/dysgeusia; symptomatic: anosmia/hyposmia or ageusia/dysgeusia or more than 2 symptoms including fever, chills, sore throat, cough, dyspnoea, diarrhoea, nausea/vomiting, headache, fatigue, and myalgia 95% CI: 95% confidence interval; SARS-CoV-2: severe acute respiratory syndrome coronavirus 2

Among the infected, 27·6% sought medical care and most received it. A small minority (1·9%) was hospitalized for over 24 hours, 13·3% were told they were suspected of having COVID-19, 4·3% performed an RT-PCR for SARS-CoV-2, and 13·5% performed a point of care test/serology for SARS-CoV-2 (Table 5).

**Table 5.**
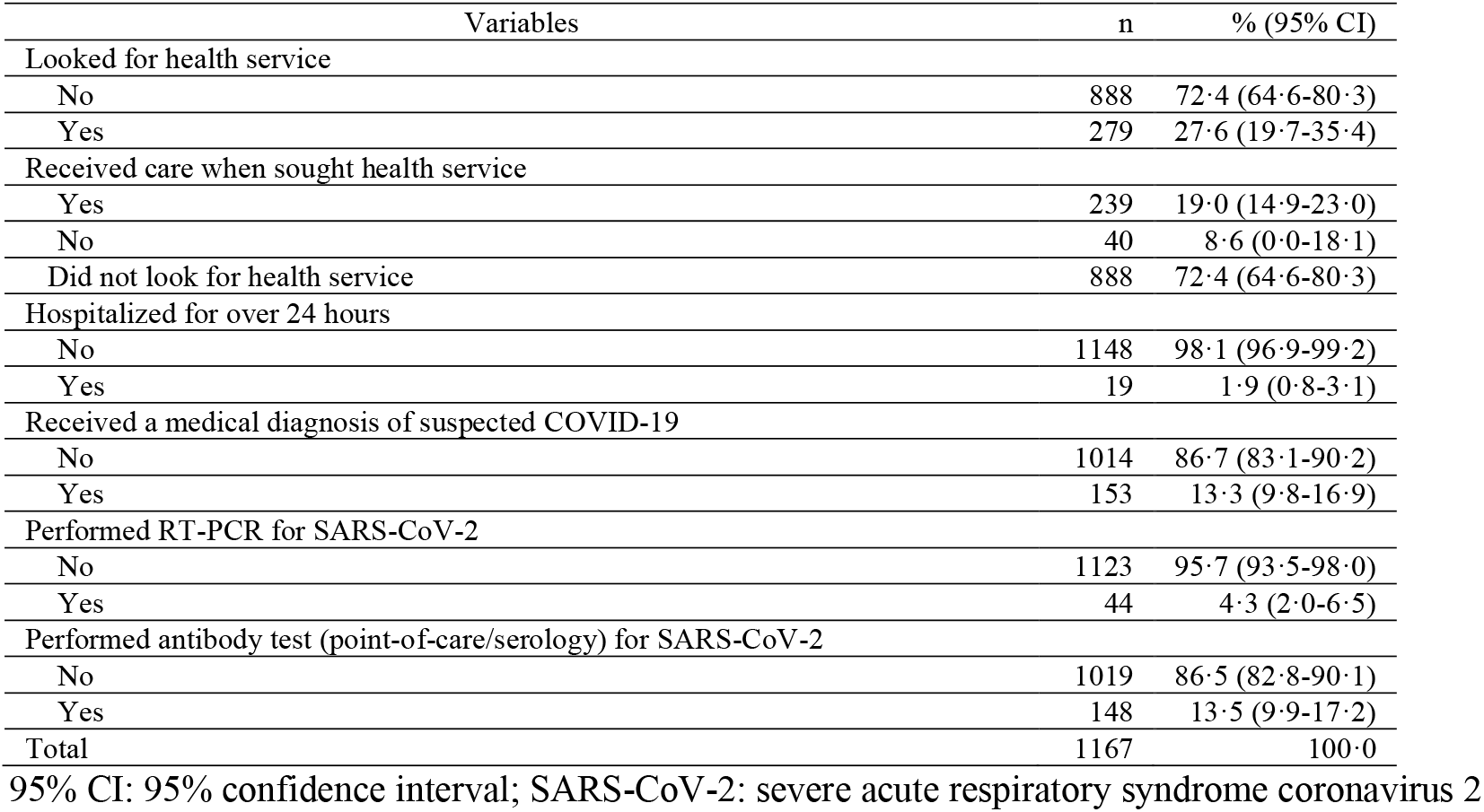
Use of health services by individuals with SARS-CoV-2 antibodies, State of Maranhão, Brazil, 2020.

The IFR was 0·15% for the State of Maranhão, and 0·30% for the São Luís Island taking reporting delays by NobBS into account. The estimates increased a little to 0·17% and 0·34%, respectively, after allowing underreporting of COVID-19 deaths. IFR was higher for males and advanced age groups (Table 6). The case reporting rate was 4·5% for the State of Maranhão, and 3·4% for the São Luís Island, resulting in a ratio of the estimated infection to the reported cases as 22·2 for the State of Maranhão, and 29·9 for the São Luís Island (Table 7).

**Table 6.** Estimated number of infections, deaths, and infection fatality rates of SARS-CoV-2 by sex and age groups, State of Maranhão and São Luís Island, Brazil, 2020.

| Sex | Age Group, years | Estimated number of infections | Number of Deaths (estimated by nowcasting) | IFR (%) | Number of Deaths (estimated by nowcasting and accounting for 15% underreporting) | IFR (%) |
| --- | --- | --- | --- | --- | --- | --- |
| Male |  |  |  |  |  |  |
|  | 0-9 | 274,241 | 9 | 0.00 | 11 | 0.00 |
|  | 10-19 | 236,559 | 13 | 0.01 | 15 | 0.01 |
|  | 20-29 | 303,111 | 22 | 0.01 | 26 | 0.01 |
|  | 30-39 | 198,478 | 76 | 0.04 | 90 | 0.05 |
|  | 40-49 | 100,908 | 163 | 0.16 | 192 | 0.19 |
|  | 50-59 | 101,695 | 280 | 0.28 | 329 | 0.32 |
|  | 60-69 | 64,637 | 623 | 0.96 | 733 | 1.13 |
|  | ≥70 | 67,000 | 1437 | 2.15 | 1691 | 2.52 |
|  | Total | 1,299,992 | 2624 | 0.20 | 3087 | 0.24 |
| Female |  |  |  |  |  |  |
|  | 0-9 | 228,212 | 16 | 0.01 | 18 | 0.01 |
|  | 10-19 | 323,171 | 5 | 0.00 | 6 | 0.00 |
|  | 20-29 | 320,049 | 18 | 0.01 | 21 | 0.01 |
|  | 30-39 | 282,436 | 62 | 0.02 | 73 | 0.03 |
|  | 40-49 | 155,868 | 83 | 0.05 | 98 | 0.06 |
|  | 50-59 | 132,770 | 174 | 0.13 | 204 | 0.15 |
|  | 60-69 | 94,033 | 359 | 0.38 | 422 | 0.45 |
|  | ≥70 | 48,838 | 932 | 1.91 | 1097 | 2.25 |
|  | Total | 1,533,005 | 1648 | 0.11 | 1939 | 0.13 |
| Overall |  |  |  |  |  |  |
|  | 0-9 | 500,448 | 25 | 0.00 | 29 | 0.01 |
|  | 10-19 | 567,266 | 18 | 0.00 | 21 | 0.00 |
|  | 20-29 | 628,088 | 40 | 0.01 | 47 | 0.01 |
|  | 30-39 | 505,975 | 139 | 0.03 | 163 | 0.03 |
|  | 40-49 | 275,270 | 246 | 0.09 | 289 | 0.11 |
|  | 50-59 | 237,395 | 453 | 0.19 | 533 | 0.22 |
|  | 60-69 | 162,429 | 982 | 0.60 | 1155 | 0.71 |
|  | ≥70 | 116,065 | 2370 | 2.04 | 2788 | 2.40 |
|  | Total | 2,877,454 | 4272 | 0.15 | 5026 | 0.17 |
| São Luís Island |  |  |  |  |  |  |
| Overall | Total | 556,611 | 1695 | 0.30 | 1883 | 0.34 |

SARS-CoV-2: severe acute respiratory syndrome coronavirus 2; IFR: infection fatality rate

**Table 7.** Estimated number of infections, number of official reported cases, case reporting rate, and ratio of the estimated infections to the reported cases of SARS-CoV-2, State of Maranhão and São Luís Island, Brazil, 2020.

| Estimates | Maranhão | São Luís Island |
| --- | --- | --- |
| Estimated number of infections (95% CI) | 2,877,454 | 556,611 |
| Number of officially reported cases | 129,700 | 18,646 |
| Case reporting rate % (95% CI) | 4.5 | 3.4 |
| Ratio of the estimated infections to the reported cases (95% CI) | 22.2 | 29.9 |

95% CI: 95% confidence interval; SARS-CoV-2: severe acute respiratory syndrome coronavirus 2

Figures 1 and 2 show dates of introduction of compulsory NPI, the weekly number of deaths by dates of occurrence and reporting, and estimates of the weekly number of deaths based on NobBS taking reporting delays, and reporting delays plus underreporting into account. The pandemic peaked from 17 May 2020 to 23 May 2020 in the State of Maranhão and from 3 May 2020 to 9 May 2020 in the São Luís Island. Since then, the number of deaths has been decreasing, and economic activity has been gradually increasing while most restrictions, apart from banning mass gatherings and opening of public schools and universities, have been eased. Nearly three months since the beginning of the relaxation of social distancing, and despite increasing community mobility, reported deaths analysed by date of occurrence remain low.

**Figure 1.**
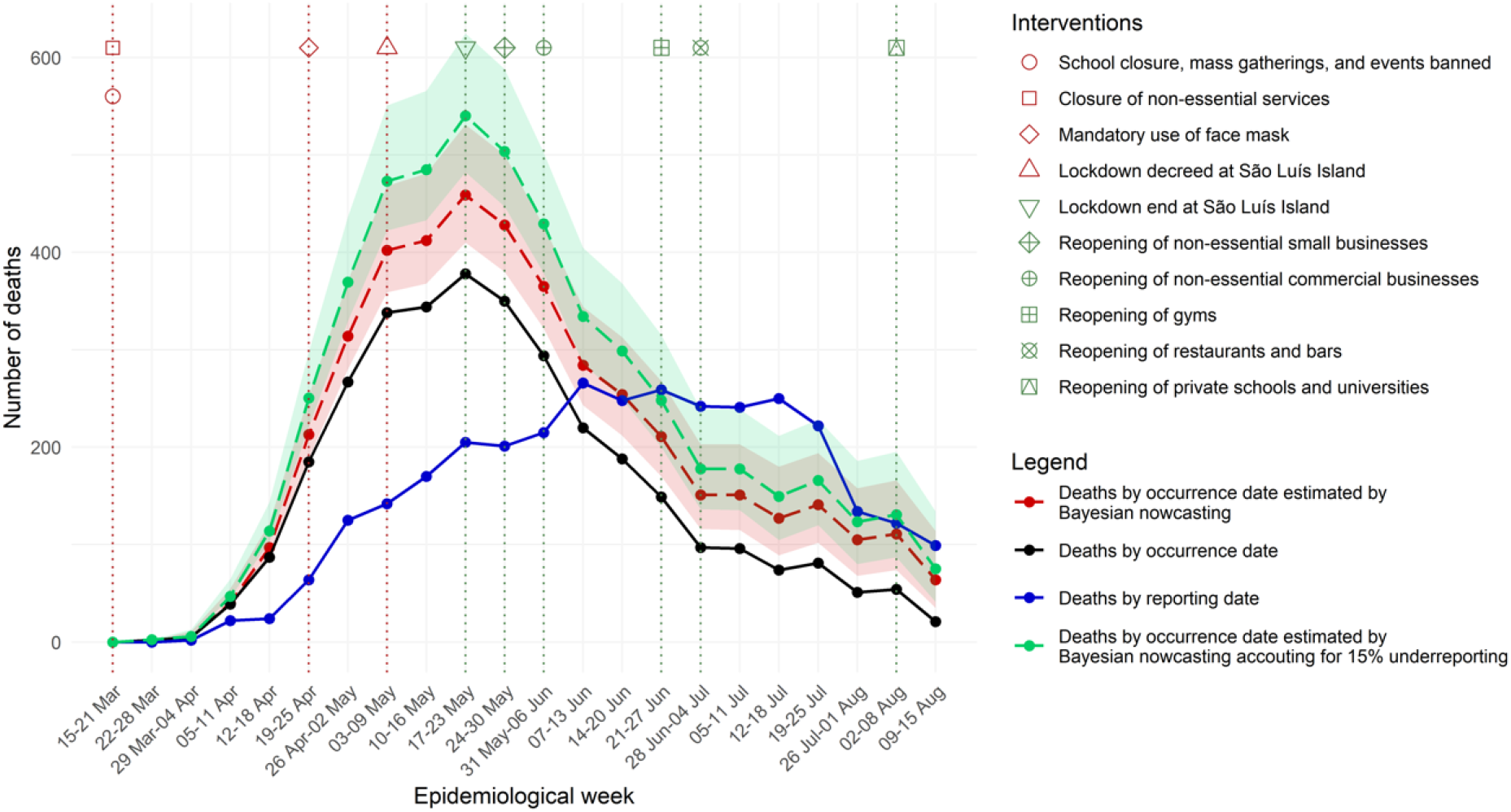
Weekly number of deaths by occurrence and reporting date, and estimated by Bayesian nowcasting from 15 March to 15 August, State of Maranhão, Brazil, 2020.

**Figure 2.**
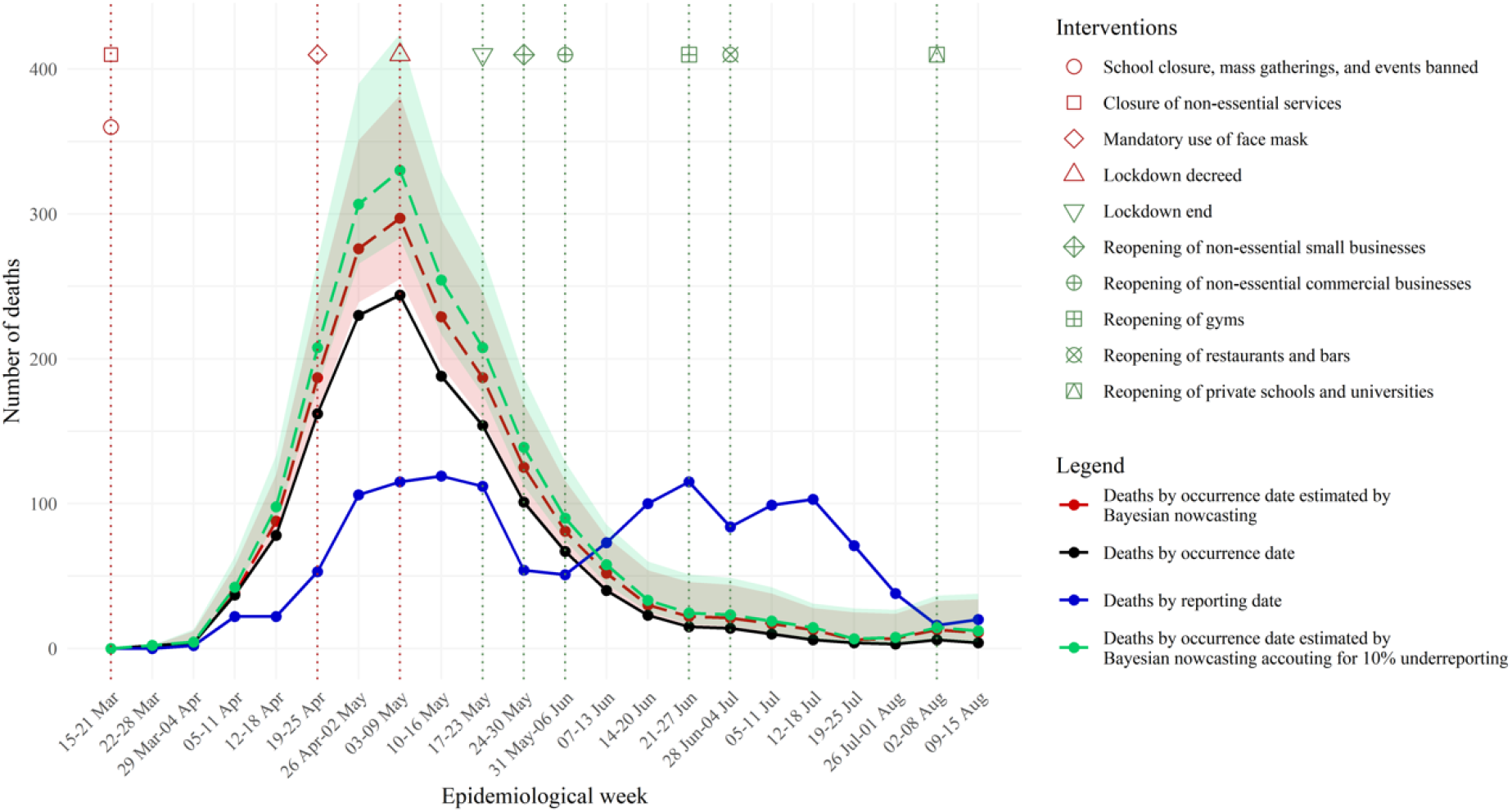
Weekly number of deaths by occurrence and reporting date, and estimated by Bayesian nowcasting from 15 March to 15 August, São Luís Island, State of Maranhão, Brazil, 2020.

## Discussion

Population-based seroprevalence of SARS-CoV-2 in the State of Maranhão, Brazil was 40·4%. To the best of our knowledge, this is the first population-based study to report a prevalence rate in this range, for an area as big as Italy. Results suggest that Maranhão is more than halfway through the herd immunity threshold, whatever its value. Although there is still doubt about what this threshold is, our data suggest that it is at least ≥40% and it does not seem to be as low as 20% as it has been suggested by some.^6^ However, herd immunity may not be uniform across populations, because it depends on whether the mixing of individuals across populations is homogeneous or heterogeneous and on the immunity duration.^6^

It seems that over 90% of all infected people develop detectable antibodies against SARS-CoV-2 two weeks after infection.^27^ In addition, SARS-CoV-2 leads to robust memory T cell responses, suggesting that infection may at least prevent subsequent severe disease.^28^ Furthermore, cross-reactivity between SARS-CoV-2 and coronaviruses that cause the common cold may elicit additional protection against infection.^29^ Due to all these factors and based on a high seroprevalence of 40·4% achieved in the survey, our data suggests that herd immunity may be achieved sooner than expected.

Nevertheless, the achievement of herd immunity will not be sustained if protection wanes over time.^30^ Thus, durable immunity may not be attained before vaccination, and consequently, the population would remain susceptible to future recurrent outbreaks.^6^

In this study, we used the Elecsys® Anti-SARS-CoV-2 electrochemiluminescence immunoassay, which presented a high specificity rate of 99·7% (95%CI 99·2-100·0) and a positive predictive value (PPV) of 97·4% with a 10% seroprevalence rate.^31^ We have likely achieved a higher PPV because the prevalence in our survey was four times that seroprevalence rate. It has been shown that electrochemiluminescence immunoassays present higher sensitivity than lateral flow immunoassays.^22^ Most existing lateral flow immunoassays do not attain an ideal performance to be used in seroprevalence surveys, especially if they are used with finger-prick.^21^ Therefore, as the test we used is more sensitive and specific, we were able to detect a higher percentage of persons with antibodies against SARS-CoV-2 with few false-positive results. The distribution and percentage of self-reported symptoms among the infected in our survey were similar to what has been reported by others,^9,11,32^ providing further evidence that a high false-positive rate in our study is unlikely.

Most surveys did not report the difference in the infection rates by sex.^9,11,13^ Some surveys have shown that adolescents^10^ and those aged between 20-59 years tend to have higher infection rates.^9,10^ We did not find evidence proving that infection rates differ by sex, age group, skin colour, or income, but our sample size given the survey’s complex sampling design lacked the statistical power to answer these questions. The infection rates were lower among those with tertiary education, in agreement with the São Paulo study.^33^

Infection rates were lower among mask wearers and among those who maintained social and physical distancing, suggesting that the use of face masks^34^ and social^35,36^ and physical distancing^34^ were necessary to avert further infections and deaths. However, adherence to NPI to curb the COVID-19 pandemic tended to diminish over time.

Infected persons were mostly symptomatic (62·2%), and anosmia/hyposmia and ageusia/dysgeusia were the two most reported symptoms. Most cases were mild. These findings are in agreement with recent studies.^18,32^

Our estimate of the IFR for the State of Maranhão was lower than the rate (0·71%) estimated for Brazil,^9^ the 0·90% estimate described for the UK and the combined estimate of 0·68% from a meta-analysis by Meyerowitz-Katz et al. (2020),^17^ but more in line with the 0·24% combined estimate obtained by Ioannidis (2020)^10^ and with the range of 0·30%-0·50% estimated by Bayesian Network Analysis.^15^ It has been shown that variations in IFR may be due to differences in the testing capacity, age structures, selective testing of high-risk populations, patterns of how deaths are attributed to COVID-19,^6^ and strain on the health services.^37^ Therefore, IFR is likely to vary across populations. However, the IFR in Maranhão is one of the lowest reported to date,^10^ even after taking reporting delays and underreporting of COVID-19 deaths into account. The BCG vaccination that is widely used in Brazil may be one of the reasons to explain the lower IFR.^38^ Another possibility is that the circulating strain in Maranhão is more transmissible and less virulent.^39^ The more rapid and extensive spread of SARS-CoV-2 in the São Luís Island may be one of the reasons to explain its higher IFR than the entire State of Maranhão.

In our study, the case reporting rate was 4·5% for the State of Maranhão and 3·4% for the São Luís Island, resulting in a ratio of the estimated infection to the reported cases as 22·2 for the State of Maranhão and 29·9 for the São Luís Island. These ratios were higher than the value of 10·3 reported for Brazil.^9^

Our study has strong points: it is population-based, had a high response rate of 77·4%, and we used a serum testing electrochemiluminescence immunoassay instead of a lateral flow immunoassay with finger-prick. There are some limitations: for some estimates, the confidence intervals were wide, and thus our power to detect statistically significant associations was lower than desired; some population groups (males and people of working age) were underrepresented in our sample.

## Contributors

Antônio A M Silva, Lídio G Lima-Neto, Conceição M P S Azevedo, Léa M Costa, Maylla L B M Bragança, Allan K D B Filho, Bernardo B Wittlin, Bruno L C A Oliveira, Carolina A Carvalho, Erika B A F Thomaz, Eudes A Simões-Neto, Jamesson F Leite-Júnior, Marcos A G Campos, Rejane C S Queiroz, Vitória A Carvalho, Vanda M F Simões, Maria T S B Alves and Alcione M Santos contributed to the conception and design of the work, the acquisition, analysis, and interpretation of the data, and the draft of the manuscript. Bruno F Souza, Sérgio S Costa and Lécia M S Cosme contributed to the acquisition and analysis of the data. All authors have approved the submitted version.

## Data Availability

We can share the data if requested by the journal.

## Acknowledgments

The study was funded by the State Health Department of Maranhão, Brazil.

## Research in context

### Evidence before this study

Of all the countries, Brazil is one of the most severely affected by coronavirus disease 2019 (COVID-19) pandemic. The government response has been controversial, and social distancing in the country has not reached levels sufficient to curb and contain the pandemic so far. Therefore, Brazil has become one of the global hotspots of the COVID-19 pandemic. Maranhão is one of the states in Brazil where the pandemic gathered speed early. Population-based surveys are necessary to monitor infection progression since most cases are undetected. However, few population-based studies on the prevalence of severe acute respiratory syndrome coronavirus 2 (SARS-CoV-2) have been performed to date, especially in low-income and middle-income countries, and most of them have used lateral flow immunoassays with finger-prick, which may yield false-negative results, and thus underestimate the true infection rate. Therefore, population-based surveys using more sensitive diagnostic tests are warranted. In this population-based study, we estimated the overall seroprevalence of SARS-CoV-2 in the State of Maranhão, Brazil using a serum testing electrochemiluminescence immunoassay.

### Added value of this study

We documented that seroprevalence of total antibodies against SARS-CoV-2 was 40·4% (95%CI 35·6-45·3). We reported that the population adherence to non-pharmaceutical interventions was higher at the beginning of the pandemic than in the last month and that SARS-CoV-2 infection rates were significantly lower among mask wearers and among those who maintained social and physical distancing in the last month compared to their counterparts. We also showed that among the infected, 26·0% were asymptomatic and 11·1% had one or two symptoms and that the predominant symptoms among those who tested positive for SARS-CoV-2 were anosmia/hyposmia (49·5%), ageusia/dysgeusia (47·7%), fever (45·6%), headache (45·4%), myalgia (43·6%), and fatigue (41·1%). The infection fatality rate was 0.17%, higher for males and advanced age groups. Only 4·5% of all infections have been reported. The ratio of the estimated infections to the reported cases was 22·2.

### Implications of all the available evidence

The reported population-based seroprevalence of 40·4%, was the highest and the closest to the herd immunity threshold reported to date, for an area as big as Italy. Whatever that threshold, the State of Maranhão in Brazil is more than halfway through the herd immunity. Our results suggest that the herd immunity threshold is not as low as 20% but at least higher than or equal to around 40%. There are suggestions that a sizeable part of the population may not be susceptible to the SARS-CoV-2 infection due to robust memory T cell responses and/or cross-reactivity between SARS-CoV-2 and other coronaviruses that cause the common cold or due to heterogeneity in susceptibility or exposure to infection. If these suggestions are true, with a high prevalence of 40·4%, we may achieve herd immunity sooner than expected. The infection fatality rate was 0·17%, one of the lowest reported so far, even after accounting for reporting delay and underreporting of COVID-19 deaths using Bayesian nowcasting.

